# Genetically-influenced HDL concentration, composition and modulation and the risk of sepsis: a Mendelian Randomization Study

**DOI:** 10.1101/2025.03.31.25324957

**Authors:** Mohan Li, Sander Kooijman, Ko Willems van Dijk, John J.P. Kastelein, Patrick C.N. Rensen, Raymond Noordam

**Affiliations:** Department of Internal Medicine, Division of Endocrinology, and Einthoven Laboratory of Experimental Vascular Medicine, Leiden University Medical Center, Leiden, the Netherlands; Department of Human Genetics, Leiden University Medical Center, Leiden, the Netherlands; NewAmsterdam Pharma B.V., Naarden, the Netherlands; Department of Vascular Medicine, Academic Medical Center, University of Amsterdam, Amsterdam, the Netherlands; Department of Clinical Epidemiology, Leiden University Medical Center, Leiden, the Netherlands; Health Campus the Hague/Public Health and Primary Care, Leiden University Medical Center, the Hague, the Netherlands

**Author notes:** Corresponding Author: Raymond Noordam PhD, Department of Clinical Epidemiology, Leiden University Medical Center, Leiden, the Netherlands, Albinusdreef 2, 2333 ZA Leiden, the Netherlands. **Competing Interests and Funding:** The authors declare no competing interests or external funding. **Author contributions:** ML conception of the work, analysis and interpretation of data, drafting the work; SK, PCNR, RN conception of the work, interpretation of data, and critical review of the work; KWD, SA, JJPK interpretation of data, and critical review of the work.

**Keywords:** High density lipoprotein, sepsis, apolipoprotein A1, cholesteryl ester transport protein, Mendelian randomization

## Abstract

**Purpose:** A growing body of evidence suggests a protective role for high-density lipoprotein (HDL) in the development and progression of sepsis. However, evidence favoring causality between HDL and sepsis remains limited. Here we investigated, by using Mendelian randomization (MR), the potential causal link between HDL particle concentration or HDL-related proteins, and the risk of developing sepsis.

**Methods:** Two-sample MR analyses were conducted to investigate association between genetically-influenced HDL particle concentration, HDL-related proteins, and the risk of sepsis. We utilized a genome-wide association study (GWAS) comprising 10,154 sepsis cases and 452,764 controls as outcome. Inverse-variance weighted (IVW) analyses were employed as primary analyses. Protein-protein interaction (PPI) network analyses were used to identify key regulators for concentration of HDL particles and ApoA1.

**Results:** Based on the IVW analyses, we found evidence for an association between genetically-influenced higher HDL particle concentration and lower risk of sepsis (odds ratio (OR): 0.86, 95% confidential interval (CI): 0.77-0.96). Similarly, genetically-influenced higher ApoA1 concentration was associated with a lower risk of sepsis (OR: 0.90, 95% CI: 0.85 - 0.96). Through PPI network analyses, we identified cholesteryl ester transfer protein (CETP) as one of the key players in regulating the concentration of HDL particles and ApoA1, and sepsis risk as such.

**Conclusion:** Our MR study provides evidence for an inverse relationship between HDL particle concentration or ApoA1, and the susceptibility to sepsis. We anticipate that CETP inhibitory strategies may increase HDL particle concentration and apoA1 to lower sepsis susceptibility and potentially prevent the risks associated with sepsis.

## Introduction

Sepsis is a life-threatening (multi-) organ dysfunction resulting from a excessive host response to infection or trauma [1]. Every year, millions of individuals are affected and up to 26% of those treated for sepsis in the hospital or ICU do not survive [2]. Antibiotics are the main treatment for sepsis but do not directly suppress the dysregulated host response. Immunosuppressive mediation is generally only used when sepsis progresses to septic shock as it also interferes with concurrent anti-inflammatory responses of the host and may hamper the body’s fight against the infection.

Besides strongly elevated cytokine levels, known as the cytokine storm, pronounced changes in lipid levels can be detected in the blood of septic patients. Sepsis typically leads to an increase in levels of free fatty acids (FFA) and triglycerides (TG), accompanied by a substantial decrease in low-density lipoprotein (LDL)-cholesterol (LDL-C) and high-density lipoprotein (HDL)-cholesterol (HDL-C) [3, 4]. In particular this latter observation appears critical, as apart from its role in reverse cholesterol transport, HDL is known to bind and neutralize circulating endotoxins including lipopolysaccharides (LPS) and lipoteichoic acid (LTA), attenuate inflammatory responses and steer immune response to clear the infection [5, 6]. Most strikingly, normalization of HDL-C levels after the initial drop are a better predictor of survival than lactate, hemoglobin, creatinine, platelets, and leukocyte count [7].

A recent Mendelian randomization (MR) study, which used genetic variants associated with the exposure as instrumental variables [8], found that genetically-influenced higher levels of HDL-C were associated with a lower risk of sepsis [9]. However, when additionally accounting for the (indirect) effects on TG and LDL-C levels by conducting multivariable-adjusted MR analysis, HDL-C level was no longer associated with sepsis or sepsis-related 28-day mortality [9]. Likewise, another recent study reported associations between measured HDL-C and sepsis, but not between genetically influenced HDL-C and sepsis [10].

HDL consists of a continuum of particles that differ in number, size, shape, lipid composition, and apolipoprotein content [11]. As a result, plasma HDL-C is only a rough indicator of HDL particle concentration. Therefore, we here tested the hypothesis that HDL particle concentration and levels of apolipoproteins associated with HDL are causally related with sepsis risk. In addition, we aimed to assess associations between proteins involved in HDL metabolism and sepsis risk, to identify potential therapeutic targets for sepsis.

## Methods

### Selection of the genetic instruments

Details of the contributing GWAS datasets are listed in **Table 1** and **Table S1**. GWAS summary data including plasma particle concentrations of HDL, LDL and very low density lipoprotein (VLDL) [12], and apolipoprotein A1 (ApoA1) and apolipoprotein B (ApoB) concentrations [13] used for the present study were extracted from the Integrative Epidemiology Unit (IEU) database (https://gwas.mrcieu.ac.uk/), which is a public database containing summary-level data from over 42,000 GWAS summary datasets.

**Table 1.** Overview of Genome-Wide Association Studies utilized in this study.

| Trait | Study | Cohorts | Phenotype definition | Sample size<br>(case/control) |
| --- | --- | --- | --- | --- |
| **Exposure** |  |  |  |  |
| HDL particle concentration | Richardson et al [12], 2022 | UK Biobank | Plasma HDL particle concentration | 115,082 |
| LDL particle concentration | Richardson et al [12], 2022 | UK Biobank | Plasma LDL particle concentration | 115,082 |
| VLDL particle concentration | Richardson et al [12], 2022 | UK Biobank | Plasma VLDL particle concentration | 115,082 |
| LDL cholesterol | Willer et al [28], 2013 | Global Lipids Genetics Consortium | Plasma LDL cholesterol level | 173,082 |
| Triglycerides | Willer et al [28], 2013 | Global Lipids Genetics Consortium | Plasma triglycerides level | 177,861 |
| ApoA1 concentration | Barton et al [13], 2021 | UK Biobank | Serum apolipoprotein A1 concentration | 398,508 |
| ApoB concentration | Barton et al [13], 2021 | UK Biobank | Serum apolipoprotein B concentration | 435,744 |
| Plasma proteomics | Sun et al [15], 2023 | UK Biobank | Plasma proteomics | 54,219 |
| CETP concentration | Blauw et al [16], 2018 | Netherlands Epidemiology of Obesity | CETP serum concentration | 4,248 |
| **Outcome** |  |  |  |  |
| Sepsis | Ponsford et al [19], 2020 | UK Biobank | Cases: explicit sepsis diagnosis codes<br>Control: not diagnosed as a case | Cases: n = 10,154<br>Control: n = 454,764 |

HDL is synthesized with ApoA1 and apolipoprotein A2 (ApoA2), and additionally carries apolipoprotein C1 (ApoC1), apolipoprotein D (ApoD), apolipoprotein E (ApoE), apolipoprotein F (ApoF), apolipoprotein L1 (ApoL1) and apolipoprotein M (ApoM) [14]. Genetic instruments for plasma apolipoproteins were extracted from large-scale proteomic GWAS summary data for 2,923 proteins of 54,219 individuals from UK Biobank participants [15]. Genetic instruments for serum cholesteryl ester transfer protein (CETP) concentration were extracted from a GWAS performed in the Netherlands Epidemiology of Obesity (NEO) study [16].

The single nucleotide polymorphisms (SNPs) used in the two-sample MR were employed to ensure their genome-wide significance (P1<15×10^−8^) with exposure and independence from one another (r^2^1<10.001, kb1=110,000) after clumping to minimize linkage disequilibrium (LD). For multivariable adjustment, SNPs that demonstrated a strong association (P1<15×10^−8^) with one of the traits were used as a group of SNPs for the three exposures. SNPs exhibiting a high LD (r^2^1>10.001) were excluded. The size of the F statistic (Beta^2^/SE^2^) was calculated for each SNP, as presented in **Table S2**. An F value exceeding 10 is indicative of sufficient instrument strength [17]. Moreover, lipid data were used to identify genetic variants in drug target genes as instrumental variables for drug-target MR analysis.

SNPs located within and near the *ABCA1*, *AKT1, AP O*, *CBETP*, *GALNT2, LIPC, LIPG*, an*L*d*P L SCARB1* genes, that were strongly associated with HDL particle or ApoA1 concentration, can serve as proxies the genetically-influenced effects of these genes on HDL particle or ApoA1 concentration. To select SNPs as proxies for lipid-regulating drugs in the drug-target-MR, we first screened for SNPs that were significantly related to HDL particle or ApoA1 concentration (P1<15×10^−8^). Among these SNPs, those located close (±100 kb) to the target genes (*ABCA1*, chromosome 9: 107543287-107690436; *AKT1*, chromosome 14: 105235686-105262085; *APOB*, chromosome 2: 21224301-21266945; *CETP*, chromosome 16: 56995762-57017757; *GALNT2*, chromosome 1: 230193536-230417868; *LIPC*, chromosome 15: 58724190-58862043; *L I P*, *G*chromosome 18: 47088427-47125555; *LPL*, chromosome 8: 19796764-19824770; *SCARB1*, chromosome 12: 125261402-125348410). Additionally, SNPs with a high LD were removed (r^2^1>10.3). This screening process has been described previously [18].

In this study, we utilized a sepsis GWAS [19] comprised of 10,154 sepsis cases and 452,764 controls from the UK Biobank as outcome for all MR analyses.

### Protein-protein interaction network analyses

Protein-protein interaction (PPI) networks are composed of proteins that interact with each other to contribute in various aspects of biological processes such as signaling, regulation of gene expression, energy and material metabolism, and cell cycle regulation. To investigate regulation of genetically-influenced HDL particle and ApoA1 concentrations, PPI network analyses were utilized to identify the key regulator(s) of each.

SNPs associated with HDL particle and ApoA1 concentrations were mapped to different genes by using the PhenoScanner (www.phenoscanner.medschl.cam.ac.uk). The search tool for the Retrieval of Interacting Genes (STRING) database (https://string-db.org) [20] provides information on each predicted PPI as well as experimentally confirmed data. The version 11.0 of STRING was employed to seek for the PPI data of the mapped genes, with the species limited to “Homo sapiens” and a median confidence of 0.4. We removed the protein nodes with no interactions with other proteins. In the PPI network, the genes served as the nodes and the edges represented the associated interactions. The connectivity degree of each node, which indicates the number of interactions of the corresponding gene, were calculated by R package “visNetwork” and the results were visualized by using R package “igraph.” Genes exhibiting a level of connectivity surpassing the average were chosen as hub genes and were marked with color red.

### Statistical analysis

Methods for MR analyses of summary-level data based on two study samples have been described in detail previously [21].

After harmonization of the SNP-exposure and SNP-outcome associations by effect allele, the inverse-variance weighted (IVW) method was used as the primary method of analysis [22]. However, this method assumes that there is no directional pleiotropy and all SNPs included as instruments are valid. For this reason, we additionally performed MR Egger analysis [23] and weighted median analysis [24]. The Wald ratio method was applied if the MR estimate contained only a single SNP. Cochran’s Q test was used to assess the SNP heterogeneity [25]. MR Egger intercept method was employed to evaluate possible directional pleiotropy, as indicated by a P-value of less than 0.05 [26].

For Mediated-MR, we initially performed a two-step two-sample MR analysis involving the exposure, outcome, and potential mediator variables [27]. Based on the principle of causal Mediated-MR analysis, the relationship between exposure and outcome is expected, and the mediator is anticipated to be associated with both the exposure and the outcome, after controlling for the exposure by using multivariable-adjusted MR [27]. The following steps involve employing the product of coefficients method by utilizing results derived from two distinct MR analyses conducted separately between the exposure and mediator (Beta_1_, SE_1_) and the mediator and outcome (Beta_2_, SE_2_). Through this approach, we assessed the indirect effect (Beta = Beta_1_* Beta_2_, SE = sqrt(Beta_1_^2^ * se_2_^2^ + beta_2_^2^ * se_1_^2^ + se_1_^2^ * se_1_^2^)) and evaluated its statistical significance [27]. The direct effects were subsequently determined by subtracting the calculated indirect effect (established using the product of coefficient method) from the total effect, which was obtained from the two-sample MR analysis.

All data analyses were conducted using R software version 4.3.1 (The R Project, https://www.r-project.org/).

## Results

### HDL particle concentration is associated with the risk of sepsis

A total of 65 SNPs associated with total HDL particle concentration were utilized to conduct MR analyses (**Table S2**). Based on IVW, genetically-influenced higher total HDL particle concentration was associated with a lower risk of sepsis (OR: 0.86, 95% CI: 0.77 – 0.96, **Figure 1**; **Table S3**), whereas no associations were observed between concentrations of total LDL particles (OR: 0.98, 95% CI: 0.91 – 1.06, **Figure 1**; **Table S4**) or total VLDL particles (OR: 1.06, 95% CI: 0.98 – 1.14, **Figure 1**; **Table S5**) and the risk of sepsis.

**Figure 1.**
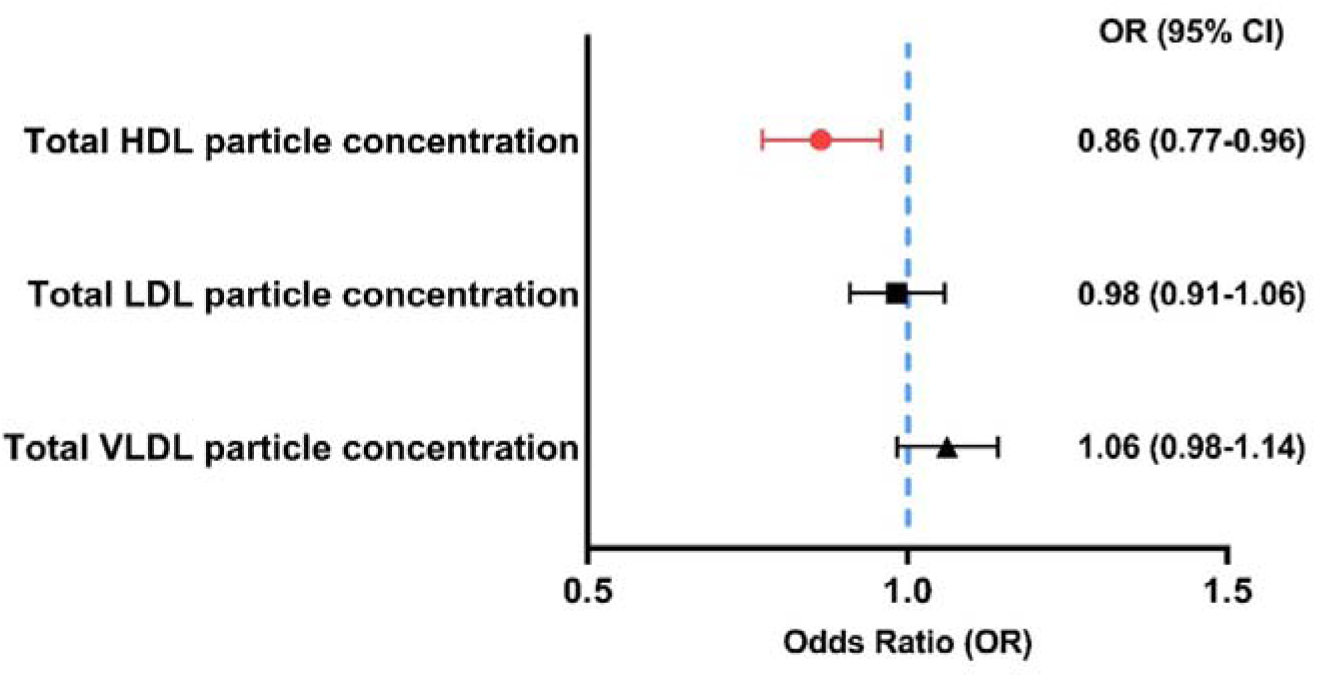
Genetically-influenced higher HDL particle concentration is associated with lower risk of sepsis. Results of two-sample Mendelian randomization between total VLDL, LDL and HDL particle concentration and risk of sepsis are shown as odds ratios (OR) with 95% confidence intervals (CI).

Similar to the univariable MR analyses, also multivariable-adjusted MR analyses showed that genetically-influenced higher total HDL particle concentration was associated with a lower risk of sepsis (OR: 0.87, 95% CI: 0.78 – 0.96, **Table S6**) after adjustment for LDL and VLDL particle concentration. Again, no association was found between LDL or VLDL particle concentration and the risk of sepsis (**Table S6**).

Additionally, multivariable-adjusted MR analyses revealed that genetically-influenced higher total HDL particle concentration was associated with a lower susceptibility after adjustment for LDL-C and TG levels (OR: 0.85, 95% CI: 0.75 – 0.96, **Table S7**).

### The possible causal association between relative plasma ApoA1 and ApoF concentration and the risk of sepsis

Since HDL particle concentration highly correlates with ApoA1 concentration, next a total of 279 SNPs associated with ApoA1 concentration were employed for MR analyses (**Table S2**). The results showed that a higher genetically-influenced ApoA1 concentration was associated with lower sepsis risk (OR: 0.90, 95% CI: 0.85 – 0.96, **Figure 2A**; **Table S8**). In contrast, genetically-influenced ApoB concentration was not associated with the risk of sepsis (OR: 1.00, 95% CI: 0.95 – 1.06, **Figure 2A**; **Table S9**).

**Figure 2.**
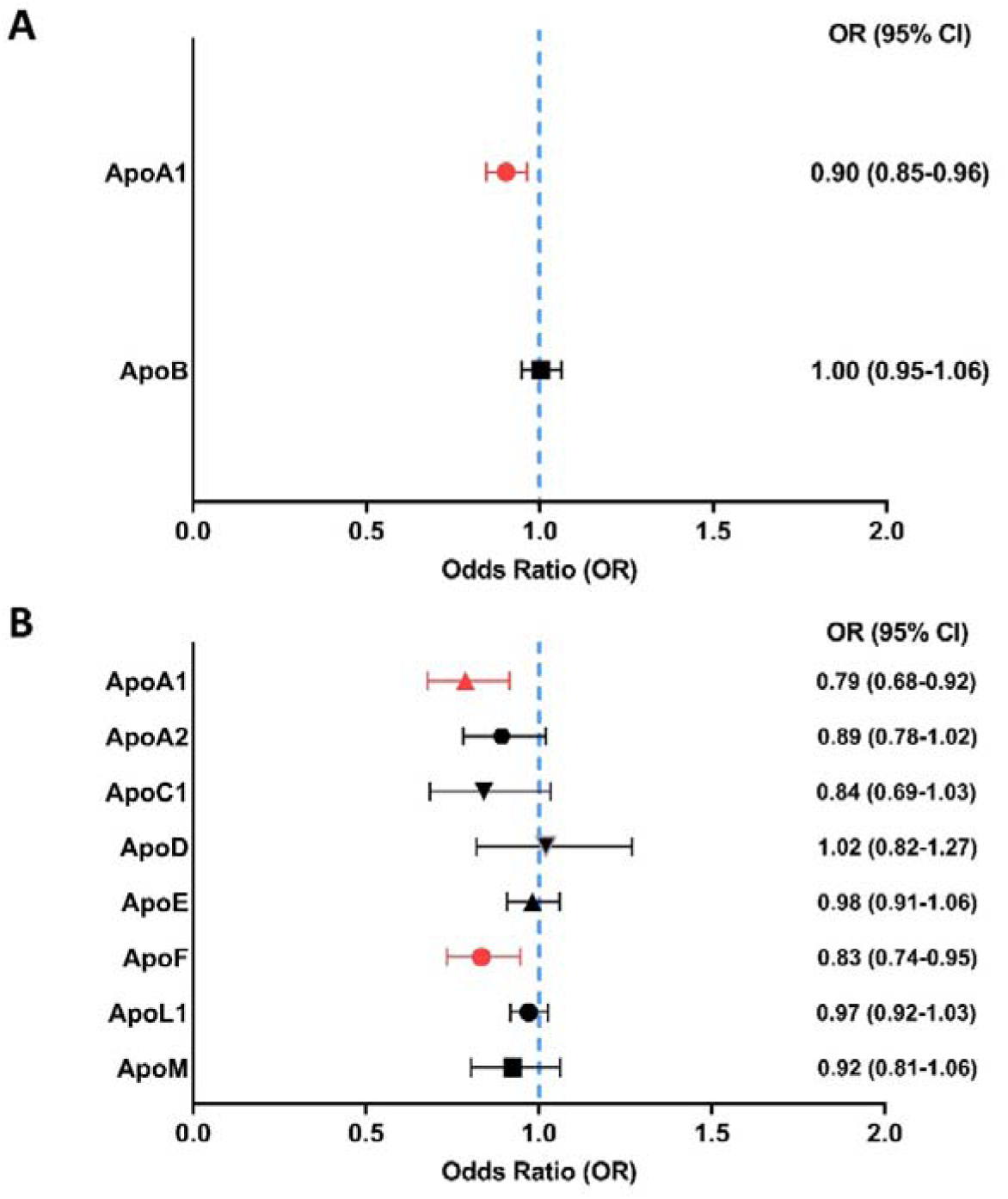
Genetically-influenced higher ApoA1 and ApoF concentration is associated with lower risk of sepsis. (A) Results of two-sample Mendelian randomization between total ApoA1 and ApoB concentrations and risk of sepsis are shown as odds ratios (OR) with 95% confidence intervals (CI). (B) Results of two-sample Mendelian randomization between sepsis and ApoA1, ApoA2, ApoC1, ApoD, ApoE, ApoF, ApoL1 and ApoM are shown as OR with 95% CI.

Subsequently, using the genetic instruments for HDL-related apolipoproteins (**Table S2**) derived from large-scale plasma proteomics dataset of the UK Biobank, we confirmed that higher genetically-influenced ApoA1 concentration was associated with lower sepsis risk (OR: 0.79, 95% CI: 0.68 – 0.92, **Figure 2B**; **Table S10**). In addition, genetically-influenced higher plasma ApoF concentration was associated with lower risk of sepsis (OR: 0.83, 95% CI: 0.74 – 0.95, **Figure 2B**; **Table S10**). This is of interest because ApoF serves as a natural inhibitor of CETP and its presence modifies the activity of CETP, resulting in reduced transfer of cholesterol from HDL to the LDL [29].

### Hub genes for regulating concentrations of HDL particles and ApoA1 were identified based on protein-protein interaction information

A total of 65 independent SNPs, mapped to 49 genes, exhibited significant associations with the total HDL particle concentration (**Table S11**). The STRING tool was employed to determine protein-protein interaction among these genes (**Table S12**), revealing *ABCA1*, *AKT1, ANGPTL4, APOB*, *APOC1, APOC4, CETP*, *GALNT2, LCAT*, *LIPC*, *LIPG*, *LPL*, and *SCARB1* as potential hub genes of genetically-influenced HDL particle concentration (**Figure 3**).

**Figure 3.**
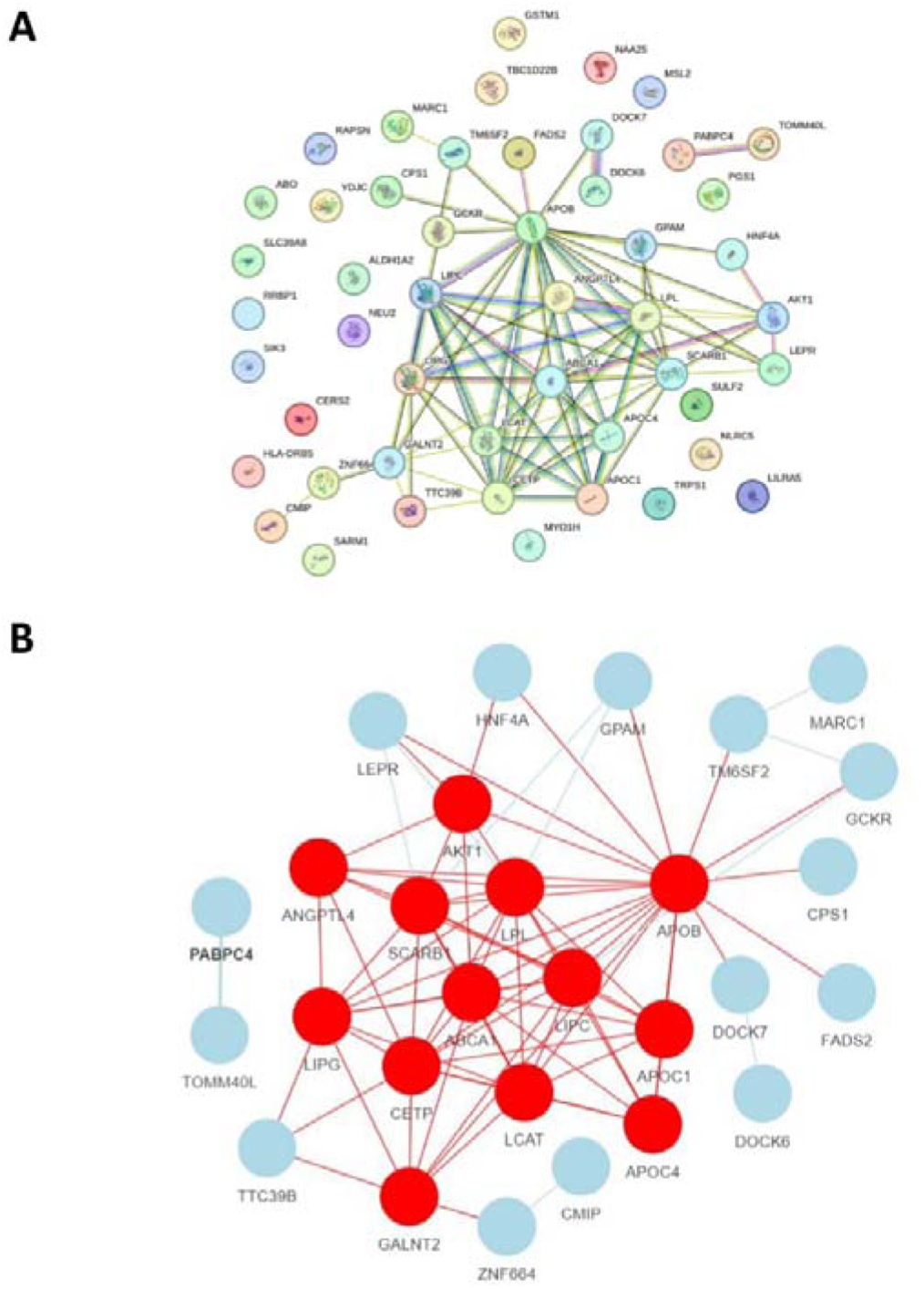
Protein-protein interaction network of genes related with total HDL particle concentration. (A) Protein-protein interaction of genes associated with total HDL particle concentration. (B) Genes identified as hub genes, which exhibit connectivity levels above the average connectivity with other genes, are highlighted in red.

By using a similar procedure, 279 SNPs, mapped to 216 genes, were found to be significantly associated with ApoA1 concentration (**Figure 4**; **Table S13**). Again, the STRING tool was used to study potential protein-protein interaction (**Table S14**), revealing 45 potential hub genes of genetically-influenced ApoA1 concentration and *ABCA1*, *AKT1*, *APOB*, *APOE*, *CD36*, *CETP*, *HSPA4*, *LIPC* and *SCARB1* as top 10 hub genes (**Figure 4**).

**Figure 4.**
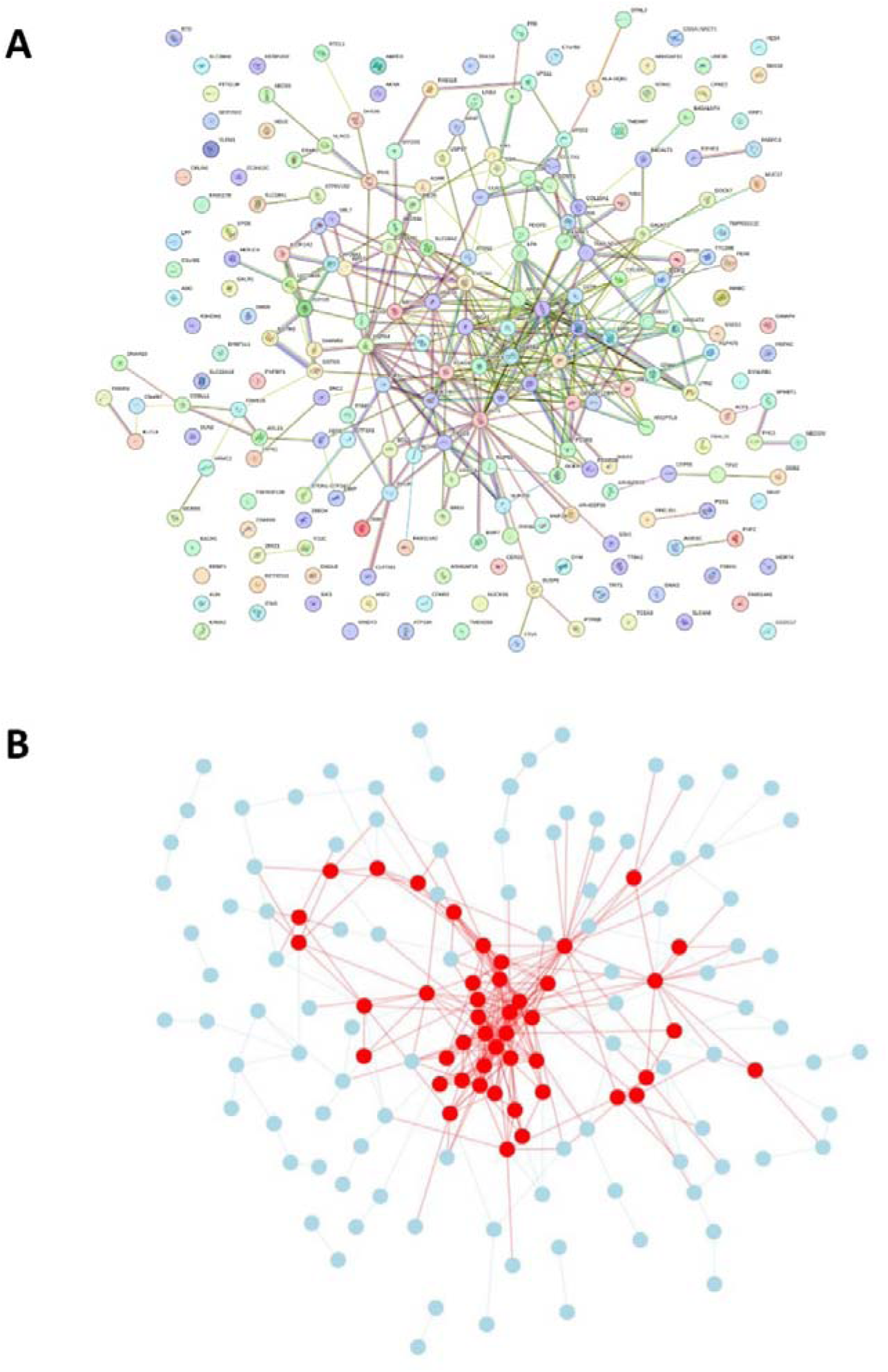
Protein-protein interaction network of genes related with ApoA1 concentration. (A) Protein-protein interaction of genes associated with ApoA1 concentration. (B) Genes identified as hub genes, which exhibit connectivity levels above the average connectivity with other genes, are highlighted in red.

We continued with studying whether the hub proteins might be a therapeutic target for mitigating the risk of sepsis. For this we used the genes that appeared in hubs for both HDL particle concentration and ApoA1 concentration, which included *ABCA1*, *AKT1, AP O*, *BCETP*, *GALNT2, LIPC, LIPG, LPL* and *SCARB1*.

### ABCA1 and CETP may serve as therapeutic targets to increase HDL particle and ApoA1 concentration thus preventing the risk of sepsis

To explore the potential of the hub genes/proteins as targets for drugs that regulates HDL particle concentration, we employed two-sample MR to examine the relationship between the effects of targeting at *ABCA1*, *A K T*, *1A P O*, *BC E T*, *PG A L N*, *TL2IPC, LIPG*, *L P L*and *SCARB1* on genetically-influenced HDL particle concentration and the risk of sepsis (**Figure 5A**). We identified 9 SNPs for *ABCA1*, 1 SNPs for *AKT1*, 5 SNPs for *APOB*, 44 SNPs for *CETP*, 3 SNPs for *GALNT2*, 12 SNPs for *LIPC*, 28 SNPs for *LIPG*, 20 SNPs for *LPL*, and 2 SNPs for *SCARB1* that serve as genetic proxies for HDL particle concentration (**Table S15**). And we identified 29 SNPs for *ABCA1*, 6 SNPs for *AKT1*, 13 SNPs for *APOB*, 57 SNPs for *CETP*, 15 SNPs for *GALNT2*, 44 SNPs for *LIPC*, 26 SNPs for *LIPG*, 23 SNPs for *LPL*, and 17 SNPs for *SCARB1* that serve as genetic proxies for ApoA1 concentration (**Table S17**).

**Figure 5.**
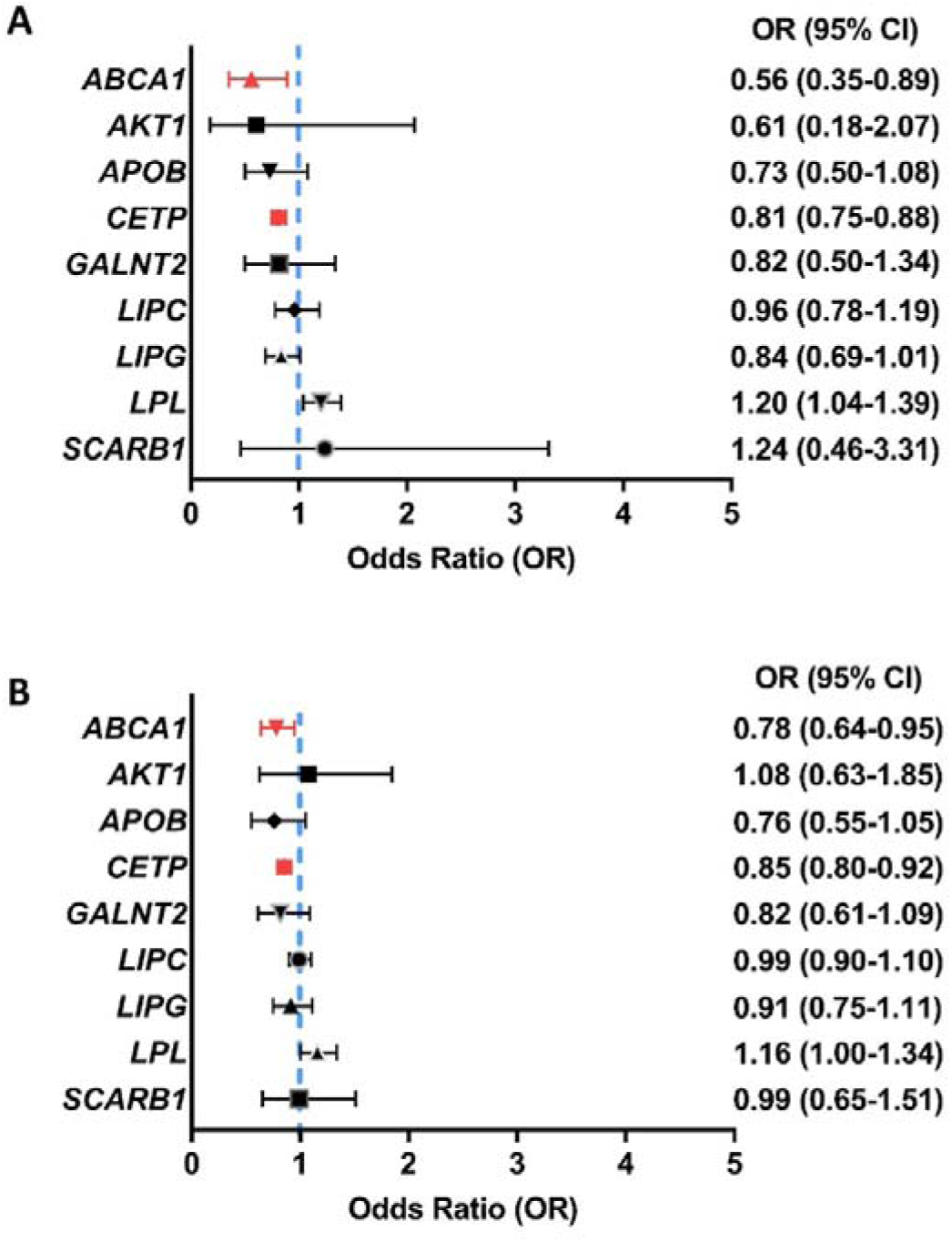
Genetically-influenced HDL particle concentration regulating and ApoA1 concentration-regulating genes and the risk of sepsis. GWAS on HDL particle and ApoA1 concentration were used as source of instrumental variants of the HDL particle concentration and ApoA1 concentration-regulating genes ABCA1, AKT 1, APO B, CET P, GALN T,2LIPC, LIPG, LPL and SCARB1. (A) Results of genetically regulating HDL particle and (B) ApoA1 concentration via ABCA1, AKT1, APOB, CETP, GALNT2, LIPC, LIPG, LPL and SCARB1 and the risk of sepsis are shown as odds ratios (OR) with 95% confidence intervals.

Targeting at *ABCA1* (OR: 0.56, 95% CI: 0.35 – 0.89, **Figure 5A**; **Table S16**) and *CETP* (OR: 0.81, 95% CI: 0.75 – 0.88, **Figure 5A**; **Table S16**) to regulate HDL particle concentration were shown to be association with lower risk of sepsis. Similarly, genetically-influenced higher ApoA1 via targeting at *ABCA1* (OR: 0.78, 95% CI: 0.64 – 0.95, **Figure 5B**; **Table S18**) as well as *CETP* (OR: 0.85, 95% CI: 0.80 – 0.92, **Figure 5B**; **Table S18**) genes were associated with a lower risk of sepsis.

### Genetically-influenced serum CETP concentration is associated with HDL particle concentration, ApoA1 concentration, and the risk of sepsis

To the best of our current knowledge, there is not yet a pharmacological approach available to directly enhance the function of ATP-binding cassette transporter A1 (ABCA1) at present. However, small molecule inhibitors targeting CETP have been developed and demonstrated effectiveness in raising plasma HDL-C levels, along with increasing concentration of ApoA1, as evidenced in a recent phase 2 randomized clinical trial [30, 31]. Both drug-target MR analyses revealed CETP as a significant common target, suggesting that targeting CETP to increase HDL particle concentration and ApoA1 levels was associated with a reduced risk of sepsis. Subsequently, we employed MR analyses to further explore whether targeting CETP could be a viable therapeutic approach for mitigating the risk of sepsis.

By using 3 SNPs associated with serum CETP concentration [16] as instrumental variables, we showed that genetically-influenced higher CETP concentration was associated with lower concentration of total HDL particles (beta: −0.48 SD, 95%CI: −0.50 to −0.47, **Table S19**) as well as lower concentration of ApoA1 (beta: −0.61 SD, 95%CI: −0.67 to −0.56, **Table S20**). Conversely, genetically-influenced higher CETP concentration was associated with a higher risk of sepsis (OR: 1.12, 95% CI: 1.05 – 1.20, **Table S21**).

We next conducted mediation analyses to investigate whether the effect of CETP serum concentration on the risk of sepsis was mediated by HDL particle concentration or ApoA1 concentration. First, multivariable-adjusted MR analyses were employed to assess whether HDL particle concentration (**Table S22**) or ApoA1 concentration (**Table S23**) was independently associated with the risk of sepsis. Only ApoA1 concentration remained associated with the risk of sepsis after being adjusted for serum CETP concentration (OR: 0.91, 95% CI: 0.83 – 0.99). We conclude that ApoA1 was shown to mediate the association between CETP serum concentration and the risk of sepsis with an effect of the mediator of 0.063 (P = 0.0026). Since the overall effect of serum CETP concentration on the risk of sepsis was 0.117 (**Table S21**), and the relative mediator effect was 53.5% (0.063/0.117). Nearly half of the impact of serum CETP concentration on sepsis risk (46.5%) could be attributed to CETP’s effects beyond its role in regulating ApoA1 levels.

## Discussion

In the present study, we found evidence for a causal association between HDL particle concentration as well as ApoA1 concentration, and the risk of sepsis. We identified CETP as a druggable key player in mitigating risk of sepsis, which at least in part works through modulating ApoA1 levels.

Previous research identified an inverse relationship between measured HDL-C levels and risk of sepsis [10]. However, a recent MR study indicated that this relationship may be confounded by LDL-C and TG levels [9]. Interesting enough, confounding effects of LDL-C and TG levels have also been observed in the association between HDL-C levels and cardiovascular disease [32]. However, even after being adjusted for HDL-C, LDL-C and TG levels, genetically-influenced higher HDL particle concentration remains associated with reduced risk of coronary artery disease [33]. Thus we hypothesized that HDL particle concentration might also play a potentially causal role in the protection from sepsis, and the rationale for this is that plasma HDL can bind with endotoxin resulting in neutralization and clearance of endotoxin [34]. Furthermore, the components of HDL, including lipids and proteins, also play a role in neutralizing LPS. Specifically, ApoA1 alone has the capability to neutralize LPS, and alterations in the structure of ApoA1 can affect this interaction [35]. HDL particles are selectively remodeled whereby specifically medium- and small-sized HDL particles are lost and larger HDL particles increase in number during inflammatory states [36, 37]. Studies comparing the predictive values of HDL-C levels and HDL particle concentration suggest that a reduced HDL particle concentration is a superior parameter over HDL-C levels [38–40]. In this study, we observed that higher genetically-influenced HDL particle concentration was associated with lower risk of sepsis before and after adjustment for LDL and VLDL particle concentration. Even after adjusting for LDL-C and TG levels through multivariable-adjusted MR analyses, the robustness of this link persisted, indicating that the relationship between HDL particle concentration and sepsis is unlikely to be confounded by LDL-C and TG levels.

The profound association between genetically-influenced ApoA1 concentration and the susceptibility to sepsis identified in this study suggests a potentially prominent role for ApoA1 in preventing sepsis. A crucial aspect of ApoA1’s function is its contribution to the anti-inflammatory effects associated with HDL, since ApoA1 is capable of mitigating the secretion of cytokines induced by LPS [41, 42]. Mice lacking ApoA-I exhibited elevated levels of serum interleukin-6 (IL-6) and tumor necrosis factor-α (TNF-α). These mice also displayed impaired neutralization and clearance of LPS, potentially amplifying TLR4/NF-κB activation [43]. In line with this, these ApoA-I-knockout mice had an increased susceptibility to sepsis-related mortality [43, 44]. Given the LPS-neutralizing capabilities of HDL and ApoA1, it was hypothesized that reconstituted HDL, formed from phospholipids and ApoA1 [45–47] and ApoA1 mimetic peptides could serve as a valuable therapeutic agent in sepsis treatment [48–50]. Intriguingly, a phase-2a randomized clinical trial including patients with sepsis showed that a synthetic HDL composed of a human recombinant ApoA-I and phospholipids, CER-001, significantly reduced lipopolysaccharides concentration in circulation [51].

Based on protein-protein interaction information, we identified CETP as one of the key regulators for both genetically-influenced HDL particle concentration and ApoA1 concentration. In line with this, lower genetically-influenced CETP serum concentration was associated with higher HDL particle concentration as well as ApoA1 concentration. This aligns with prior clinical trials that demonstrated CETP inhibitors effectively elevate HDL particle [52] and ApoA1 [53, 54] concentration. CETP facilitates the exchange of cholesteryl esters with TGs between HDL and TG-rich lipoproteins. This process is followed by hepatic lipase (HL)-mediated breakdown of the TG in the HDL, leading to the formation of lipid-poor ApoA1, which is subsequently eliminated through renal filtration, ultimately reducing the number of HDL particles [55]. Vice versa, pharmacologically inhibiting CETP may increase the lipidation of ApoA1, thus slowing down its renal clearance and potentially increasing HDL particle as well as ApoA1 concentration. Previous clinical observational studies have shown that higher levels and activity of CETP during the early stages of sepsis are related with future organ damage and mortality [56–58]. In addition, a rare missense variant in CETP (rs1800777-A) is linked to a significant reduction in HDL-C levels during the occurrence of sepsis and carriers of the A allele had lower 28-day survival rates compared to non-carriers [56], demonstrating the significance of CETP in the pathogenesis and progression of sepsis. Likewise, we found that genetically-influenced lower CETP serum concentration was associated with a lower risk of sepsis. Interestingly, our research also showed an association between elevated genetically-influenced plasma ApoF concentration and a lower risk of sepsis, in line with the observation that ApoF serves as a natural CETP inhibitor [29]. By using APOE*3-Leiden.CETP mice, a model for human lipoprotein metabolism, Trinder et al. found that mice pre-treated with the CETP inhibitor anacetrapib for three weeks displayed significantly higher HDL-C levels as well as less severe symptoms of endotoxemia after cecal ligation and puncture than mice treated with placebo [59]. Likewise, administrating anacetrapib after the onset of cecal ligation and puncture-induced sepsis significantly lowered plasma creatinine levels and improved survival rates [59]. Intriguingly, our mediation MR results revealed that targeting at the regulation of CETP serum levels might not solely decrease sepsis risk via elevating ApoA1 concentration. Instead, CETP serum concentration also appears to independently influence susceptibility to sepsis. Taken together, these results further support the idea that CETP may play a vital role in the pathogenesis, complications, and the severity of sepsis, thus pharmacologically inhibiting CETP might be a plausible strategy for preventing and improving the outcome of sepsis.

We should carefully interpret the results because of the inherent limitations of MR studies, including the assumption of instrument validity, potential population stratification, and pleiotropy of genetic variants. The present study was conducted in European-ancestry individuals only, thus extrapolating the findings to non-European populations should be done with caution. Furthermore, the question remains whether individuals with genetically-influenced higher concentration of HDL particles or ApoA1 under normal circumstances could maintain relatively elevated levels or experience faster normalization during infection or sepsis. It is also important to acknowledge that the genetically-influenced serum concentration of CETP may not completely represent the effects of pharmacologically inhibiting CETP activity. Consequently, there remains a necessity for relevant pre-clinical and clinical studies to address these matters.

In conclusion, this MR study indicated that genetically-influenced higher concentration of HDL particles and ApoA1 are associated with a lower risk of sepsis. Our findings point to a potentially causal role of elevated CETP serum concentration in the pathogenesis of sepsis and highlight the importance of pharmacologically inhibiting CETP strategies to prevent the susceptibility to sepsis.

## Supplementary materials

**Table S1** The different experimental groups carried out in this study

**Table S2** SNPs for lipids in Two-sample MR

**Table S3** Results of MR between concentration of HDL particles and sepsis

**Table S4** Results of MR between concentration of LDL particles and sepsis

**Table S5** Results of MR between concentration of VLDL particles and sepsis

**Table S6** SNPs for concentration of lipoprotein particles in multivariable-adjusted MR (concentration of HDL, LDL, VLDL particles)

**Table S7** SNPs for concentration of lipoprotein particles in multivariable-adjusted MR (concentration of HDL particle, LDL-C, TG levels)

**Table S8** Results of MR between ApoA1 concentration and sepsis

**Table S9** Results of MR between ApoB concentration and sepsis

**Table S10** Results of MR between plasma apolipoprotein (proteomics) concentration and sepsis

**Table S11** Mapped genes of concentration of HDL particles

**Table S12** PPI between mapped genes of concentration of HDL particles

**Table S13** Mapped genes of ApoA1 concentration

**Table S14** PPI between mapped genes of ApoA1 concentration

**Table S15** SNPs for HDL particle concentration-regulating targets in drug-target MR

**Table S16** Result of MR between HDL particle concentration-regulating targets and sepsis

**Table S17** SNPs for ApoA1-regulating targets in drug-target MR

**Table S18** Result of MR between ApoA1 concentration-regulating targets and sepsis

**Table S19** Results of MR between CETP serum concentration and concentration of HDL particles

**Table S20** Results of MR between CETP serum concentration and ApoA1 concentration

**Table S21** Results of MR between CETP serum concentration and sepsis

**Table S22** SNPs for concentration of lipoprotein particles and CETP serum concentration in multivariable-adjusted MR

**Table S23** SNPs for ApoA1 concentration and CETP serum concentration in multivariable-adjusted MR

## Data Availability

All data produced in the present study are available upon reasonable request to the authors

## References

1. Singer, M., et al., The Third International Consensus Definitions for Sepsis and Septic Shock (Sepsis-3). Jama, 2016. 315(8): p. 801–10.

2. Fleischmann-Struzek, C., et al., Incidence and mortality of hospital-and ICU-treated sepsis: results from an updated and expanded systematic review and meta-analysis. Intensive Care Med, 2020. 46(8): p. 1552–1562.

3. Lee, S.H., et al., Prognostic Implications of Serum Lipid Metabolism over Time during Sepsis. Biomed Res Int, 2015. 2015: p. 789298.

4. Pirillo, A., A.L. Catapano, and G.D. Norata, HDL in infectious diseases and sepsis. Handb Exp Pharmacol, 2015. 224: p. 483–508.

5. Levine, D.M., et al., In vivo protection against endotoxin by plasma high density lipoprotein. Proc Natl Acad Sci U S A, 1993. 90(24): p. 12040–4.

6. Jiao, Y.L. and M.P. Wu, Apolipoprotein A-I diminishes acute lung injury and sepsis in mice induced by lipoteichoic acid. Cytokine, 2008. 43(1): p. 83–7.

7. Barber, G., J. Tanic, and A. Leligdowicz, Circulating protein and lipid markers of early sepsis diagnosis and prognosis: a scoping review. Curr Opin Lipidol, 2023. 34(2): p. 70–81.

8. Emdin, C.A., A.V. Khera, and S. Kathiresan, Mendelian Randomization. Jama, 2017. 318(19): p. 1925–1926.

9. Lou, C., et al., Genetic association of lipids and lipid-lowering drugs with sepsis: a Mendelian randomization and mediation analysisF. ront Cardiovasc Med, 2023. 10: p. 1217922.

10. Liu, G., et al., The relationship between high density lipoprotein cholesterol and sepsis: A clinical and genetic approach. Clin Transl Sci, 2023. 16(3): p. 489–501.

11. Zannis, V.I., et al., HDL biogenesis, remodeling, and catabolism. Handb Exp Pharmacol, 2015. 224: p. 53–111.

12. Richardson, T.G., et al., Characterising metabolomic signatures of lipid-modifying therapies through drug target mendelian randomisation. PLoS Biol, 2022. 20(2): p. e3001547.

13. Barton, A.R., et al., Whole-exome imputation within UK Biobank powers rare coding variant association and fine-mapping analyses. Nat Genet, 2021. 53(8): p. 1260–1269.

14. Phillips, M.C., New insights into the determination of HDL structure by apolipoproteins: Thematic review series: high density lipoprotein structure, function, and metabolism. J Lipid Res, 2013. 54(8): p. 2034–2048.

15. Sun, B.B., et al., Plasma proteomic associations with genetics and health in the UK Biobank. Nature, 2023. 622(7982): p. 329-338.

16. Blauw, L.L., et al., CETP (Cholesteryl Ester Transfer Protein) Concentration: A Genome-Wide Association Study Followed by Mendelian Randomization on Coronary Artery Disease. Circ Genom Precis Med, 2018. 11(5): p. e002034.

17. Burgess, S., S.G. Thompson, and C.C.G. Collaboration, Avoiding bias from weak instruments in Mendelian randomization studies.International journal of epidemiology, 2011. 40(3): p. 755–764.

18. Rosoff, D.B., et al., Mendelian Randomization Study of PCSK9 and HMG-CoA Reductase Inhibition and Cognitive Function. J Am Coll Cardiol, 2022. 80(7): p. 653–662.

19. Ponsford, M.J., et al., Cardiometabolic Traits, Sepsis, and Severe COVID-19: A Mendelian Randomization Investigation. Circulation, 2020. 142(18): p. 1791–1793.

20. von Mering, C., et al., STRING: a database of predicted functional associations between proteins. Nucleic Acids Res, 2003. 31(1): p. 258–61.

21. Noordam, R., et al., Assessment of causality between serum gamma-glutamyltransferase and type 2 diabetes mellitus using publicly available data: a Mendelian randomization study. International journal of epidemiology, 2016. 45(6): p. 1953–1960.

22. Burgess, S., A. Butterworth, and S.G. Thompson, Mendelian randomization analysis with multiple genetic variants using summarized dataG. enet Epidemiol, 2013. 37(7): p. 658–65.

23. Bowden, J., G. Davey Smith, and S. Burgess, Mendelian randomization with invalid instruments: effect estimation and bias detection through Egger regression. Int J Epidemiol, 2015. 44(2): p. 512–25.

24. Bowden, J., et al., Consistent estimation in Mendelian randomization with some invalid instruments using a weighted median estimator. Genetic epidemiology, 2016. 40(4): p. 304–314.

25. Greco, M.F., et al., Detecting pleiotropy in Mendelian randomisation studies with summary data and a continuous outcome. Stat Med, 2015. 34(21): p. 2926–40.

26. Verbanck, M., et al., Detection of widespread horizontal pleiotropy in causal relationships inferred from Mendelian randomization between complex traits and diseases. Nat Genet, 2018. 50(5): p. 693–698.

27. Carter, A.R., et al., Mendelian randomisation for mediation analysis: current methods and challenges for implementation. Eur J Epidemiol, 2021. 36(5): p. 465–478.

28. Willer, C.J., et al., Discovery and refinement of loci associated with lipid levels. Nat Genet, 2013. 45(11): p. 1274–1283.

29. Liu, Y. and R.E. Morton, Apolipoprotein F-A natural inhibitor of CETP and key regulator of lipoprotein metabolism. Current opinion in lipidology, 2020. 31(4): p. 194.

30. Nicholls, S.J., et al., Lipid lowering effects of the CETP inhibitor obicetrapib in combination with high-intensity statins: a randomized phase 2 trial. Nat Med, 2022. 28(8): p. 1672–1678.

31. Ballantyne, C.M., et al., Obicetrapib plus ezetimibe as an adjunct to high-intensity statin therapy: A randomized phase 2 trial. J Clin Lipidol, 2023. 17(4): p. 491–503.

32. Holmes, M.V., et al., Mendelian randomization of blood lipids for coronary heart disease. Eur Heart J, 2015. 36(9): p. 539–50.

33. Zhao, Q., et al., A Mendelian randomization study of the role of lipoprotein subfractions in coronary artery disease. Elife, 2021. 10.

34. Emancipator, K., G. Csako, and R.J. Elin, In vitro inactivation of bacterial endotoxin by human lipoproteins and apolipoproteins. Infect Immun, 1992. 60(2): p. 596–601.

35. Wang, Y., et al., Effect of lipid-bound apoA-I cysteine mutants on lipopolysaccharide-induced endotoxemia in mice. J Lipid Res, 2008. 49(8): p. 1640–5.

36. de la Llera Moya, M., et al., Inflammation modulates human HDL composition and function in vivo. Atherosclerosis, 2012. 222(2): p. 390–4.

37. McGillicuddy, F.C., et al., Inflammation impairs reverse cholesterol transport in vivo. Circulation, 2009. 119(8): p. 1135–1145.

38. Kontush, A., HDL particle number and size as predictors of cardiovascular disease. Front Pharmacol, 2015. 6: p. 218.

39. Kuller, L.H., et al., Lipoprotein particles, insulin, adiponectin, C-reactive protein and risk of coronary heart disease among men with metabolic syndrome. Atherosclerosis, 2007. 195(1): p. 122–8.

40. Mackey, R.H., et al., High-density lipoprotein cholesterol and particle concentrations, carotid atherosclerosis, and coronary events: MESA (multi-ethnic study of atherosclerosis).J Am Coll Cardiol, 2012. 60(6): p. 508–16.

41. Emancipator, K., G. Csako, and R. Elin, In vitro inactivation of bacterial endotoxin by human lipoproteins and apolipoproteins. Infection and immunity, 1992. 60(2): p. 596–601.

42. Flegel, W., et al., Prevention of endotoxin-induced monokine release by human low-and high-density lipoproteins and by apolipoprotein AI. Infection and immunity, 1993. 61(12): p. 5140–5146.

43. Guo, L., et al., High density lipoprotein protects against polymicrobe-induced sepsis in mice. Journal of Biological chemistry, 2013. 288(25): p. 17947–17953.

44. Li, Y., J.-B. Dong, and M.-P. Wu, Human ApoA-I overexpression diminishes LPS-induced systemic inflammation and multiple organ damage in mice. European journal of pharmacology, 2008. 590(1-3): p. 417–422.

45. Casas, A.T., et al., Reconstituted high-density lipoprotein reduces LPS-stimulated TNFα. Journal of Surgical Research, 1995. 59(5): p. 544–552.

46. Tanaka, S., et al., Reconstituted high-density lipoprotein therapy improves survival in mouse models of sepsis.Anesthesiology, 2020. 132(4): p. 825–838.

47. Guo, L., et al., Replenishing HDL with synthetic HDL has multiple protective effects against sepsis in mice. Science Signaling, 2022. 15(725): p. eabl9322.

48. Zhang, Z., et al., Apolipoprotein AI mimetic peptide treatment inhibits inflammatory responses and improves survival in septic rats. American Journal of Physiology-Heart and Circulatory Physiology, 2009. 297(2): p. H866–H873.

49. Dai, L., et al., The apolipoprotein AI mimetic peptide 4F prevents defects in vascular function in endotoxemic rats. Journal of lipid research, 2010. 51(9): p. 2695–2705.

50. Moreira, R.S., et al., Apolipoprotein AI mimetic peptide 4F attenuates kidney injury, heart injury, and endothelial dysfunction in sepsis. American Journal of Physiology-Regulatory, Integrative and Comparative Physiology, 2014. 307(5): p. R514–R524.

51. Stasi, A., et al., Beneficial effects of recombinant CER-001 high-density lipoprotein infusion in sepsis: results from a bench to bedside translational research project. BMC medicine, 2023. 21(1): p. 392.

52. Krauss, R.M., et al., Changes in lipoprotein subfraction concentration and composition in healthy individuals treated with the CETP inhibitor anacetrapib. Journal of lipid research, 2012. 53(3): p. 540–547.

53. Furtado, J.D., et al., Pharmacological inhibition of CETP (cholesteryl ester transfer protein) increases HDL (high-density lipoprotein) that contains ApoC3 and other HDL subspecies associated with higher risk of coronary heart disease. Arteriosclerosis, Thrombosis, and Vascular Biology, 2022. 42(2): p. 227–237.

54. Nicholls, S.J., et al., Lipid lowering effects of the CETP inhibitor obicetrapib in combination with high-intensity statins: a randomized phase 2 trial. Nature Medicine, 2022. 28(8): p. 1672–1678.

55. von Eckardstein, A., Mulling over the odds of CETP inhibition.Eur Heart J, 2010. 31(4): p. 390–3.

56. Trinder, M., et al., Cholesteryl ester transfer protein influences high-density lipoprotein levels and survival in sepsis. American journal of respiratory and critical care medicine, 2019. 199(7): p. 854–862.

57. Genga, K.R., et al., CETP genetic variant rs1800777 (allele A) is associated with abnormally low HDL-C levels and increased risk of AKI during sepsis. Scientific Reports, 2018. 8(1): p. 16764.

58. Grion, C.M., et al., Lipoproteins and CETP levels as risk factors for severe sepsis in hospitalized patients. Eur J Clin Invest, 2010. 40(4): p. 330–8.

59. Trinder, M., et al., Inhibition of cholesteryl ester transfer protein preserves high-density lipoprotein cholesterol and improves survival in sepsis. Circulation, 2021. 143(9): p. 921–934.

